# Mendelian randomisation with coarsened exposures

**DOI:** 10.1101/2020.08.20.20178921

**Authors:** Matthew J. Tudball, Jack Bowden, Rachael A. Hughes, Amanda Ly, Marcus R. Munafò, Kate Tilling, Qingyuan Zhao, George Davey Smith

**Author notes:** Correspondence: Oakfield House, Oakfield Grove, Bristol, UK, BS8 2BN.

## Abstract

A key assumption in Mendelian randomisation is that the relationship between the genetic instruments and the outcome is fully mediated by the exposure, known as the exclusion restriction assumption. However, in epidemiological studies, the exposure is often a coarsened approximation to some latent continuous trait. For example, latent liability to schizophrenia can be thought of as underlying the binary diagnosis measure. Genetically-driven variation in the outcome can exist within categories of the exposure measurement, thus violating this assumption. We propose a framework to clarify this violation, deriving a simple expression for the resulting bias and showing that it may inflate or deflate effect estimates but will not reverse their sign. We then characterise a set of assumptions and a straight-forward method for estimating the effect of standard deviation increases in the latent exposure. Our method relies on a sensitivity parameter which can be interpreted as the genetic variance of the latent exposure. We show that this method can be applied in both the one-sample and two-sample settings. We conclude by demonstrating our method in an applied example and re-analysing two papers which are likely to suffer from this type of bias, allowing meaningful interpretation of their effect sizes.

## 1 Introduction

Mendelian randomisation proposes to use genetic variants that alter, or mirror the biological effects of, modifiable exposures to study the causal effects of such exposures on downstream outcomes. The principle underlying Mendelian randomisation is that genetic variants are randomly passed from parents to offspring at conception, resulting in a plausibly unconfounded source of variation in the exposures with which they are associated. For Mendelian randomisation estimates to inform policies or clinical practices, we must additionally assume that genetic and environmental modifiers of the exposure produce similar effects on the outcome (Davey Smith & Ebrahim, 2003). For example, Mendelian randomisation studies of pharmaceutical exposures typically use genetic variants that code for potential drug targets, assuming that similar effects would be observed if those targets were altered therapeutically (Plump & Davey Smith, 2019).

One of the crucial assumptions underlying the Mendelian randomisation approach is that the relationship between the genetic instruments and the outcome is fully mediated by the exposure, known as the exclusion restriction assumption. However, it is important to draw a distinction between the true exposure experienced by an individual and our attempt at measuring it. For practical purposes, we are often restricted to coarsened approximations which do not fully encapsulate the mechanism by which the true exposure of interest affects the outcome. Consistent with existing terminology, we define an exposure measurement as coarsened if it is a discrete measure approximating a continuous latent exposure (Marshall, 2016).

In the Mendelian randomisation context, coarsened exposures can violate the exclusion restriction assumption. If the genetic instruments are acting on a latent exposure, such as body mass index (BMI), but the measured exposure is a discretisation of it, such as obesity status, then there can exist genetically-driven variation in the true exposure within categories of the measured exposure. We could imagine that counterfactually altering some BMI-raising single nucleotide polymorphism (SNP) in an individual could result in a change in their BMI without necessarily changing their obesity status. This can be viewed a form of measurement error which opens up potential pathways from the genetic instruments to the outcome that do not pass through the exposure measure, thus violating the exclusion restriction assumption.

For example, Richardson et al. (2020) attempt to separate the effects of early and later life adiposity on disease risk. The adiposity variable is a three-category self-report measure (‘thinner’, ‘plumper’ and ‘about average’). It is reasonable to conceptualise a continuous measure of body mass (e.g. BMI) underlying this coarsened categorical measure, such that genetic variation in this latent continuous measure could occur within categories of the self-report variable. We later re-analyse Richardson et al. (2020) in Box 2 using the approach proposed in this paper. Another example is Richmond et al. (2019), who apply Mendelian randomisation to investigate the effect of sleep traits (e.g. morning preference, sleep duration) on breast cancer risk, finding large causal effects of several traits. These traits are categorical measures, for example, morning preference is measured in six categories and sleep duration is split into several groups. It is reasonable to conceptualise the true exposures on which the genetic variants are acting as latent continuous sleep traits and preferences, for which the measured exposures are discrete markers.

An important class of latent exposures we consider in this paper is disease liabilities, for which binary disease diagnosis or case status is the typical exposure measurement. There are an increasing number of Mendelian randomisation studies investigating the effects of complex diseases such as asthma, schizophrenia and attention deficit hyperactivity disorder on various outcomes (Lawn et al., 2019; Martins-Silva et al., 2019; Pasman et al., 2018; Sun et al., 2019). Complex diseases which result from the interaction of environment and multiple genetic variants are likely to affect outcomes of interest through pathways other than diagnosis, for example, severity of sub-clinical symptoms. Since genetic instruments are, in turn, likely to influence the manifestation or severity of the underlying symptoms, rather than diagnosis alone, this represents a potential violation of the exclusion restriction.

This specific violation of the exclusion restriction assumption has been raised before in both the economics and political science literatures (Angrist & Imbens, 1995; Marshall, 2016). It has also been raised briefly in the Mendelian randomisation context in Burgess & Labrecque (2018), who discuss interpretation of estimates with binary exposures. The authors recommend that findings be framed in terms of this latent exposure but note that the estimates themselves have no meaningful causal interpretation. However, it remains to explore in more detail how this bias may distort estimates and clarify how to appropriately frame estimates in terms of the latent exposure, which will depend on the unobservable relationship between the latent exposure and its coarsened measurement.

We attempt to provide these clarifications in this paper. In particular, we derive an expression for the bias and introduce a clear set of identifying assumptions under which one can estimate the causal effect of the latent exposure. We hope to allow researchers to decide whether these assumptions are plausible in the context of their study. In Section 2, we outline our technical framework, which assumes a linear single threshold model for the relationship between the latent exposure and its measurement. That is, we assume that values of the coarsened exposure are determined by whether the latent exposure is above or below some threshold, which could be individual-specific. For example, an individual is classified as obese if their BMI is above 30 and not obese otherwise. This framework also contains the Falconer (1965) liability-threshold model, which assumes that a disease occurs in an individual, or is sufficiently pronounced to be diagnosed, if a build-up of underlying liability crosses some threshold. In this model, liability is assumed to capture all genetic, shared and non-shared environmental risk factors.

In Section 3.1, we derive an expression for the bias from the naive approach of using the coarsened measure as the exposure directly. Then, in Section 3.2, we show that, if the latent exposure is standardised to have a standard deviation of one, its causal effect can be identified if we have auxiliary information on the genetic variance of the latent exposure. This may be obtained from GWAS or treated as a sensitivity parameter and varied over a plausible range of values. In the context of disease liabilities, we may use the coefficient of determination developed by Lee et al. (2012).

Section 4 provides some generalisations to this framework, in particular, allowing two-sample estimation. Section 5 provides a real data example by creating artificially dichotomised variables from the continuous BMI measure in UK Biobank. Boxes 1 and 2 present reanalyses of two papers which could be interpreted within the framework proposed in this paper (Pasman et al., 2018; Richardson et al., 2020). In Sections A and B of the Appendix, we examine the bias that can emerge when the assumptions of our framework are violated.

## 2 Framework

We begin by outlining some key notation. Suppose there is a genetic instrument *Z* ∈ ℝ, other genetic variants (e.g. pleiotropic, weak) *X* ∈ ℝ^*K*^ and an environmental risk factor *V* ∈ ℝ, where *V* is assumed to be continuously distributed with mean zero. We also assume that *Z, X* and *V* are mutually independent. We define *G* = *µ* + *αZ* + *γ′X* as the genetic share of the latent exposure and define the latent exposure itself as

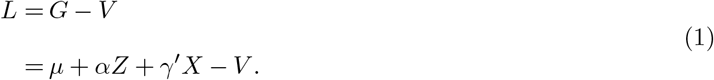

It would be equally correct to define *L* = *G* + *V*, but the formulation in (1) simplifies some later expressions. In the Falconer framework described in Section 1, *L* would represent liability to some disease. We are able to observe a coarsened exposure characterised by a dichotomisation of the latent exposure.

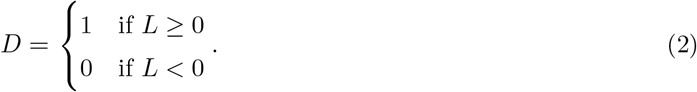

If *L* is disease liability, then *D* would represent occurrence of the disease. In practice, we measure diagnosis of the disease, which does not necessarily correspond to occurrence due to under- or over-diagnosis. We will treat the two as equivalent throughout and discuss violations of this equivalence in Section 6.

Equation (2) is the crucial assumption underlying our approach; namely, that *L* is a linear index that relates to *D* according to a single threshold. Section A of the Appendix elaborates on the importance of this structural assumption. Figure 1 illustrates our model within the Falconer framework. There is a distribution of disease liabilities and the disease occurs at the right tail of this distribution. The size of the grey region represents the prevalence of the disease in the population.

**Figure 1:**
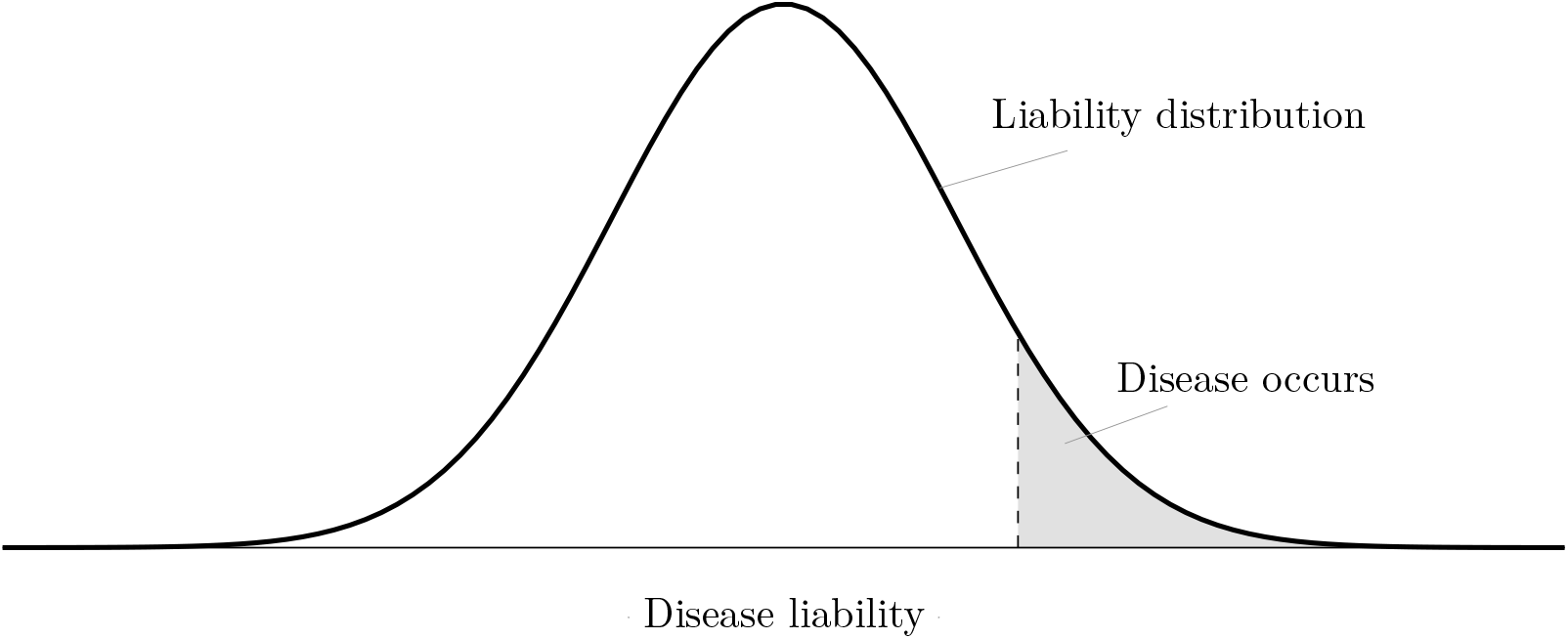
In the Falconer framework, liability to a disease is assumed to follow a smooth (often normal) distribution. The disease occurs at the tail of the distribution, with the grey region representing prevalence in the population.

We also have an observed outcome *Y* ∈ ℝ. For ease of exposition, we restrict ourselves to a simple linear structural equation model

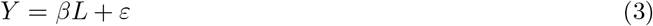

which is implicitly conditional on covariates, where *ε* can be correlated with both *V* and *X*. However, this framework can accommodate more general exposure-outcome relationships of the form *Y* = *f* (*L*) + *ε*, provided *f* (*·*) is differentiable. We make the standard instrumental variable assumptions, namely, that *α /*= 0 and *Z* is independent of *ε* conditional on covariates. The model (3) implicitly captures the assumption described in Section 1 that genetic and environmental modifiers of the exposure produce equivalent effects on the outcome. In this setting, the marginal effect (in absolute value) of both *G* and *V* is *β*. Figure 2 summarises this model in a directed acyclic graph. We can see that the exclusion restriction is violated since there exists a path from the latent exposure *L* to *Y* which does not pass through the measured exposure *D*. The structural equation (3) assumes no effect of *D* itself. For a disease such as schizophrenia, liability could have a harmful effect on the outcome but being diagnosed will usually lead to receiving treatment and thus could have a protective effect. We cannot separately identify the two effects in this setting, although possibilities for doing so are discussed in Section 4.2. When *D* is believed to have a distinct effect on the outcome, we may instead identify the total effect of liability on the outcome; i.e. the direct effect *β* and the indirect effect through *D*.

**Figure 2:**
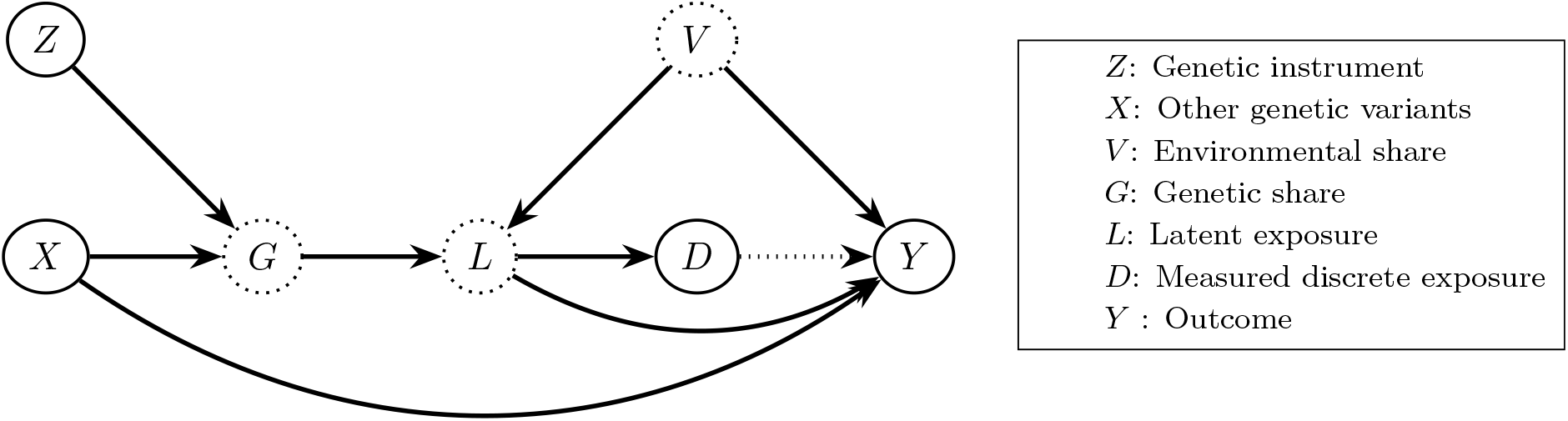
The framework proposed in Section 2 summarised in a directed acyclic graph. Dotted circles represent latent variables and complete circles represent observed variables.

The structural assumptions made in this section can be summarised as follows:

**Assumption 1**. *(Single threshold) The latent exposure L and its binary measurement D are related by a single threshold model of the form D* = *I* {*L ≥* 0}.

**Assumption 2**. *(Additivity) L* = *G* − *V, where G and V are, respectively, the genetic and environmental shares of L*.

**Assumption 3**. *(Linearity) G is a linear function of the genetic instrument Z and other genetic variants X, such that G* = *µ* + *αZ* + *γ*′*X*.

**Assumption 4**. *(Environmental share) V has mean zero, standard deviation σ*_*V*_ *and is in some family of continuous distributions, with cumulative distribution function given by F* (*v*/*σ*_*V*_) = *F*_*V*_ (*v*) *and density f* (*v*/*σ*_*V*_) = *f*_*V*_ (*v*).

**Assumption 5**. *(Risk factor independence) Z, X and V are mutually independent*.

**Assumption 6**. *(Gene-environment equivalence) The outcome model takes the form Y* = *βL* + *ε, where ε is a random disturbance and X and V may be correlated with ε*.

**Assumption 7**. *(Instrumental variable assumptions) Z is independent of ε and α /*= 0.

## 3 Identification

### 3.1 Bias from the naive approach

The naive approach to Mendelian randomisation is to use the coarsened exposure *D* as the exposure directly. We show in this section that this results in a ‘multiplicative’ bias which will scale the true effect *β* up or down, but not change its direction. When the distribution of *L* has a light tail (e.g. normal distribution), we will typically see inflation of effect estimates, with the degree of inflation increasing as the prevalence of *D* becomes smaller. If *D* is case status for a disease, for example, then effect estimates will be more inflated for rarer diseases. We see this pattern of inflation occurring in our real data examples in Section 5.

We call the naive Wald estimand *β*_*D*_ = *cov*(*Z, Y*)/*cov*(*Z, D*). It is illustrative to derive a closed-form expression for *β*_*D*_. Suppose *Z* is binary and *G* = *µ* + *αZ* (i.e. there is no *X*). Begin by noting that

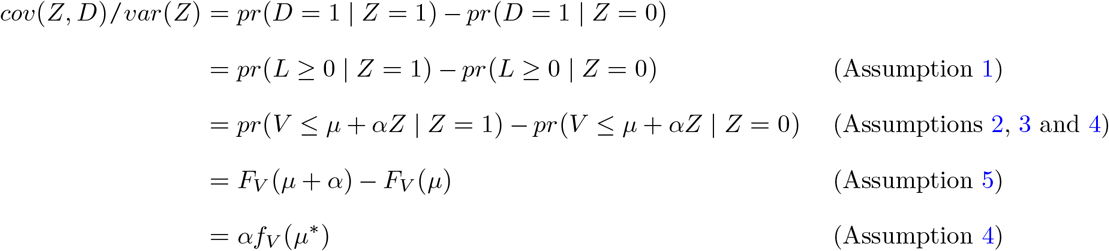

by the mean value theorem, where *µ ≤ µ*\* *≤ µ* + *α*. Thus, the estimand can be written as

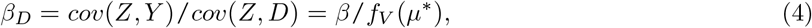

meaning that *β*_*D*_ is equal to the true latent exposure effect *β* divided by the density of *V* at the value *µ*\*. *f*_*V*_ (*µ*\*) is not identified since the distribution of *V* is unknown and *µ*\* is defined on the scale of the latent exposure.

### 3.2 The latent variable approach

The bias formula (4) indicates that the nuisance term is *f*_*V*_ (*·*), which is the distribution of the environmental share *V*. Although *D* depends on this unobserved distribution, the genetic share *G* does not. Our latent variable approach therefore proceeds in four steps: 1) estimate the linear predictor of a generalised linear model of *D* on *Z* and *X*; 2) normalise the linear predictor to have mean zero and variance one; 3) use this normalised linear predictor as the exposure in an instrumental variable model; and 4) scale the resulting effect estimate up by the genetic variance of the latent exposure. Step 4) is necessary to interpret effect estimates in terms of standard deviation increases in the latent exposure, which is typically the desired scale.

To state this more precisely, define 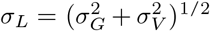 as the standard deviation of *L*, where 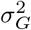 and 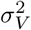 are the variances of *G* and *V* respectively. Within the framework described in Section 2, we claim that the four steps above allow us to identify *β*_*L*_ = *σ*_*L*_*β* from the observed data (*Z, X, D, Y*).

The remainder of this section proves this claim given the assumptions outlined in Section 2 and discusses its implications. We begin by expressing the quantity *pr* (*D* = 1 | *X* = *x, Z* = *z*) within the framework of Section 2.

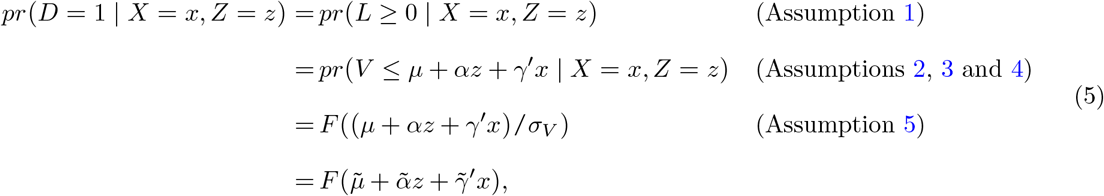

Where 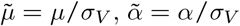 and 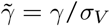. *F* can be interpreted as the link function in a generalised linear model and 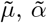 and 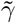 as parameters that can be identified from the observable data. In practice, we could specify *F* directly, for example, as a logistic or normal distribution (corresponding to logistic and probit regressions respectively). Alternatively, to avoid imposing potentially strong distributional assumptions, we could use semi-parametric estimation methods for generalised linear models, which only require some smoothness conditions on *F* (Ichimura, 1993; Klein & Spady, 1993). Disease liabilities are often assumed be the product of many small, independent traits. Therefore, by the central limit theorem, a normal distribution (i.e. probit model) is a natural choice of link function in this context (Curnow, 1972).

Step 1 is accomplished by constructing the predicted genetic share of the latent exposure

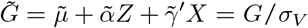

using parameters estimated from the generalised linear model of *D* on *Z* and *X*. An immediate complication is that *σ*_*V*_ is unobserved. Treating *σ*_*V*_ as a sensitivity parameter is not tractable since its value is defined on the scale of the latent exposure, which is unknown. However, if we standardise 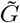 by its standard deviation as in step 2, we can remove *σ*_*V*_ since

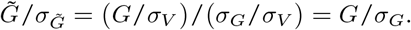

By using *G*/*σ*_*G*_ as our exposure, we can obtain effects in terms of standard deviation increases in the genetic share of the latent exposure. The instrumental variable estimand of step 3 equals

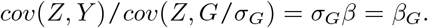

This estimand does not often have a natural interpretation. We would prefer to interpret our effects in terms of changes in the latent exposure itself.

Let 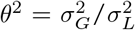 be defined as the genetic variance of the latent exposure. If we have a suitable choice of *θ*^2^, we can simply adjust our estimand as in step 4 such that

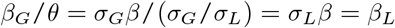

which is our desired effect. The parameter *θ*^2^ can be treated as a sensitivity parameter and varied over a plausible range of values or can, in some instances, be obtained from GWAS which report this measure.

For disease liabilities in particular, Lee et al. (2012) uses the Falconer liability-threshold model to develop a coefficient of determination for GWAS that is interpretable on the liability scale, which corresponds to *θ*^2^. Therefore, *θ*^2^ can be estimated using this approach or selected from GWAS which report this coefficient. For ease of interpretation, liability is often assumed to have mean zero and variance one, in which case *σ*_*L*_ = 1 and *β* itself is identified on this scale (Lee et al., 2012).

## 4 Some generalisations

### 4.1 Individual-specific threshold

The formalisation of the relationship between disease and liability in equation (2) and Figure 1 assumes a fixed threshold. That is, all individuals with liability above the threshold will develop or be diagnosed with the disease and all those below the threshold will not. In reality, we might imagine that diagnosis has a random component, driven, for example, by preferences of the diagnosing clinician or imprecision of the testing procedure. It might be more realistic to assume a model such that

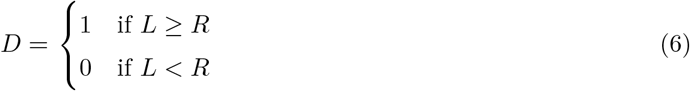

where *R* is a random individual-specific threshold. Provided *R* is independent of the instrument *Z* and other variants *X*, this random threshold will not affect identification of 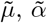 and 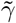 of equation (5) under correct model specification. However, the link function *F* of equation (5) no longer corresponds to the distribution family of *V*; instead, it corresponds to the distribution family of *V* + *R*. This could make correct specification of the link function more difficult and semi-parametric approaches may be warranted.

### 4.2 Identifying effects of the coarsened exposure

The structural model (3) assumes no direct effect of the binary exposure measure *D* on the outcome. As discussed in Section 3, when *D* is diagnosis of a disease, we might expect resulting treatment or therapy to have an effect on the outcome distinct from disease liability, suggesting a structural equation model of the form

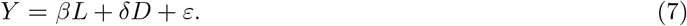

The exposure measure is downstream of the latent exposure and there are assumed to be no direct pathways from the genetic instruments to the exposure measure, as illustrated in Figure 2. Therefore, we cannot use our genetic instrument *Z* to estimate the independent effect of the exposure measure on the outcome; the genetic instruments induce no unique variation in the exposure measure independent of the latent exposure. However, consider the individual-specific threshold of Section 4.1. The variable *R* could represent preferences of the clinician for diagnosing the disease or a change in clinical practices affecting some individuals (Brookhart & Schneeweiss, 2007; Davies et al., 2013). If *R* is independent of each individual’s liability, without directly affecting the outcome, then it is a potential instrument for disease diagnosis. The general rule for separately estimating the effects of the latent exposure and dichotomisation is to have instruments which induce distinct variation in both.

### 4.3 Multi-valued discrete exposure

This method generalises easily to the multi-valued discrete exposure setting. Suppose we observe a discretised variable characterised by

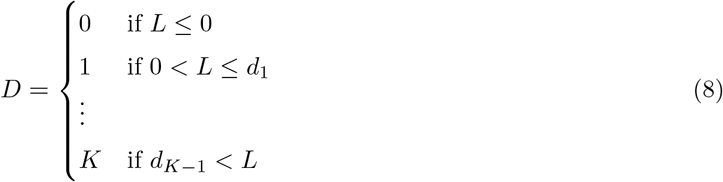

where 0 *< d*1 *<* … *< d*_*K*−_1 are latent thresholds. *D* could represent number of years in education and *L* could represent time in education as a continuous measure. Similar to how the dichotomous exposure can be formulated as a binary response model as in equation (5), exposures of the form (8) can be formulated as an ordered response model and the parameters 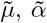 and 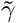 are still identified, allowing the method to be applied as usual.

### 4.4 Two-sample design with GWAS summary statistics

For rare diseases, it is not always possible to observe the coarse exposure measurement *D* and the outcome *Y* in the same sample. It is common practice in Mendelian randomisation studies to use summary statistics from separate GWAS of the exposure and outcome to obtain two-sample estimates (Burgess et al., 2015). This method also generalises to the two-sample setting using the popular inverse-variance weighted approach (Burgess et al., 2013).

Suppose there is a set 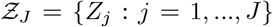 of SNPs from the exposure GWAS, of which a subset 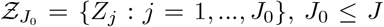, is selected as instruments from the outcome GWAS. Suppose we have estimates 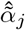 on the log-odds scale of the instrument-exposure relationship 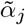 for each instrument in 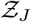 and estimates of the instrument-outcome relationship 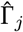for each instrument in 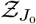. Additionally, we need the variance 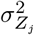 for each instrument in 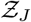, which can be obtained from reported allele frequencies. Lastly, we also need estimates for the inverse-variance weights 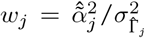, where 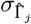 is the standard error of 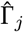. Under the assumption that the instruments in 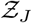 are mutually independent, the inverse-variance weighted estimator for *β*_*G*_ = *cov*(*Z, Y*)/*cov*(*Z, G*/*σ*_*G*_) can be obtained from the above summary statistics as

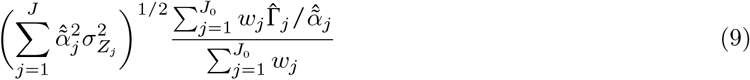

which is derived in Section C of the Appendix. We can recover the effect in terms of *σ*_*L*_ (i.e. *β*_*L*_) by rescaling by a suitable choice of *θ*^2^ as described in Section 3. Conveniently, the second term in (9) is the standard form of the inverse-variance weighted estimator. This means that we can easily readjust existing Mendelian randomisation estimates of coarsened exposures using only the exposure GWAS and a choice for *θ*^2^. The large-sample distribution of the estimator (9) is derived in Section C of the Appendix.

## 5 Real data examples

We can assess the performance of this method in a realistic setting by creating a dichotomised variable from an observed continuous measure, BMI. The idea is to dichotomise BMI at some threshold value and then treat only the dichotomisation as observed. We shall compare the true standardised effect of BMI on some outcome with our procedure described in Section 3 and with the naive approach of using the dichotomisation as the exposure.

Our example is based on the Mendelian randomisation analysis performed in Lyall et al. (2017), which estimates the effect of BMI on several cardiometabolic measures in the UK Biobank cohort. In particular, we look at the effect of BMI on systolic blood pressure. This is a convenient exposure-outcome relationship to estimate because we should not expect there to be threshold effects, i.e. the dichotomisations of BMI should have no distinct effects on systolic blood pressure except through BMI itself.

Consistent with Lyall et al. (2017), we use as potential instruments the 93 genome-wide significant SNPs reported in Locke et al. (2015) available in UK Biobank and we control for age, sex, assessment centre, alcohol intake, smoking status and Townsend deprivation index, along with genetic batch and the first 10 principal components of the genetic relatedness matrix. To avoid weak instrument bias, we prune these SNPs by including those which correlate with BMI with |*t*| *>* 4 (conditional on the other SNPs) as instruments. We estimate the ‘true’ standardised effect of BMI on systolic blood pressure via two-stage least squares, finding that a one standard deviation increase in BMI corresponds to an increase in systolic blood pressure of 1.53 mm Hg (95% CI 0.34 - 2.72). At each BMI threshold, we then generate a binary variable equal to 1 if an individual’s BMI is above the threshold and 0 otherwise. Treating only this binary measure as observed, we apply the latent variable approach of Section 3.2 using a probit link function.

The results of this example are summarised in Figure 3, which compares the estimated effects with the ‘true’ effect of 1.53. The estimates using the dichotomised measure as the exposure are highly sensitive to the choice of threshold. Since we should not expect there to be distinct threshold effects in this setting, this demonstrates that the dichotomised exposure is not capturing the effect of the latent exposure, instead, it is picking up the shape of the distribution of the environmental risk factor for BMI, as discussed in Section 3.1. As predicted by the bias formula in Section 3.1, the estimates were inflated at the extreme thresholds where the distribution is flatter.

**Figure 3:**
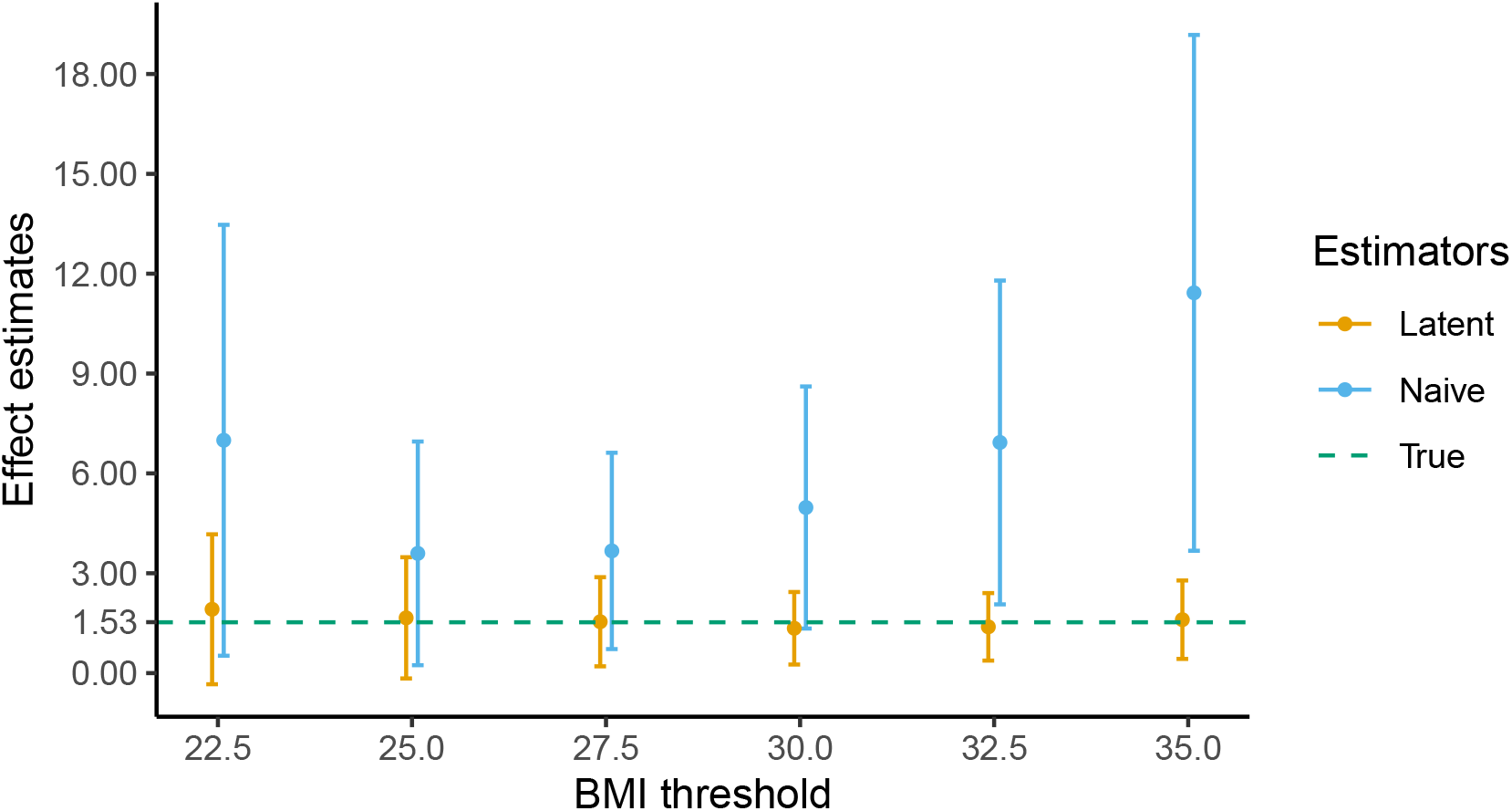
Comparison of estimated effect with ‘true’ effect for various BMI thresholds. *N* = 70,261, *θ*^2^ = 0.0256, 95% confidence intervals are generated over 1,000 bootstrap resamples. ‘True’ corresponds to the sample estimate using BMI as the exposure; ‘naive’ corresponds to using the dichotomous measurement as the exposure *β*_*D*_; and ‘latent’ corresponds to the latent variable estimator *β*_*L*_ of Section 3.2.

For the latent variable approach, we select a *θ*^2^ of 0.0256 based on the *R*-squared of our first stage regression of BMI on the genetic variants. The effect estimate from this approach is much less sensitive to the choice of threshold. Furthermore, the estimates appear to accurately recover the ‘true’ effect of 1.53 regardless of the threshold value, ranging from 1.35 at a BMI cut-off of 30 to 1.92 at a BMI cut-off of 22.5.

We can also investigate this approach in a more realistic setting by re-analysing two existing papers. Box 1 gives an example of how existing two-sample results which do not have interpretable effect sizes can be reinterpreted using this method. The original paper finds that schizophrenia liability increases one’s likelihood of using cannabis, although the effect sizes are not interpretable (Pasman et al., 2018). Using our approach, we find that a one standard deviation increase in liability corresponds to an odds ratio in the range 1.15-1.26 (95% CI 1.10-1.44) for ever using cannabis. This approach allows us to infer the size of this effect which, in this instance, is very modest.

Box 2 gives an example of how this approach can correct exclusion restriction violations. In the original paper, self-reported adiposity is measured on a three-point scale (‘thinner’, ‘plumper’ and ‘about average’). Genetic instruments will be acting on the underlying measure of child adiposity (e.g. BMI) rather than the three-point scale, so the exclusion restriction is likely to be violated (Richardson et al., 2020). We use our latent variable approach to ameliorate this bias and to estimate the effect of child BMI directly, which is the exposure of interest.

## 6 Discussion

We propose a simple framework for estimation and interpretation of Mendelian randomisation for coarsened measurements of latent continuous exposures. We begin by demonstrating in Section 3.1 that using the coarse measurement as the exposure results in a multiplicative bias which will inflate or deflate effect estimates without reversing their sign. However, under the assumptions of our framework, described in Section 2, we can recover the effect of the latent exposure in terms of standard deviation increases. Section 4.4 shows that it is straight-forward to generalise this approach to the two-sample setting. The key sensitivity parameter in our approach is the genetic share of the variance of the latent exposure, which may be estimated or varied over a plausible range of values (Lee et al., 2012). Section 5 evaluates this approach by creating binary exposure measurements from the continuous BMI measure in UK Biobank. We show that we can accurately recover the effect of a standard deviation increase in BMI on systolic blood pressure. We also demonstrate this approach in practice by re-analysing two papers which are likely to suffer from this type of exclusion restriction violation, allowing us to meaningfully interpret their effect sizes.

The approach proposed in this paper relies on a number of strong structural assumptions on the relationship between the latent exposure and its corresponding measurement. The appropriateness of these assumptions must be assessed on a case-by-case basis. Exposure measurements which are defined by strict thresholds of the latent continuous exposure are easiest to conceptualise within this framework. In general, the assumption most difficult to justify is that the thresholds are independent of the genetic share of the latent exposure. One example where this assumption may be violated is self-report measures of mental health status, for example, feelings of depression on a 1-5 scale. Individuals who are genetically predisposed to depression may have different thresholds for reporting their mental wellbeing, either over- or under-reporting.

An additional complication occurs when this method is applied to disease exposures. We have assumed throughout that disease occurrence and disease diagnosis are equivalent; that is, everyone who develops the disease will receive a diagnosis. However, there are often barriers to seeking and accessing the healthcare services needed to receive a diagnosis. These might include stigma surrounding the disease, a lack of trust in healthcare providers or a lack of access to healthcare services due to cost, distance or institutional complexities (Cassim et al., 2019; Stangl et al., 2019). It is therefore possible that individuals with the disease will fail to be diagnosed. This can be viewed as a form of misclassification bias. Misclassification-robust methods for binary exposures could potentially be incorporated into this approach, which we leave for future work (Lewbel, 2000; Rekaya et al., 2016; Smith et al., 2013).

In studies where the assumptions in Section 2 are believed to be implausible, it is important for researchers to be transparent that the magnitude of their effect estimate will not be well-defined.

**Box 1:** Re-analysis of Pasman et al. (2018)

Pasman et al. (2018) performs a two-sample bi-directional Mendelian randomisation analysis of schizophrenia and cannabis use (Burgess et al., 2015). The gene-exposure associations for schizophrenia are pulled from a GWAS of cases and controls and are reported on the log-odds scale (Schizophrenia Working Group of the Psychiatric Genomics Consortium, 2015). While this avoids the problem of using the dichotomous diagnosis variable as the exposure (as discussed in Section 1), it means that the resulting estimates are interpreted as unit increases in the log-odds, which are scaled by the unobserved parameter *σ*_*V*_. The authors report an odds ratio of 1.16 (95% CI 1.06 - 1.27) for the effect of genetic liability to schizophrenia. While we can infer the direction of the effect from this estimate, we cannot draw any conclusions about the magnitude.

We apply the two-sample generalisation of Section 4.4. One of the strengths of this generalisation is that we do not need to re-estimate the original inverse-variance weighted Mendelian randomisation estimates ourselves. In addition to the estimates reported in the original paper, we need only an estimate of *σ*_*G*_, which can be computed from summary data from the schizophrenia GWAS, and some plausible choice∗s for the sensitivity parameter *θ*^2^. The schizophrenia GWAS reports that their genome-wide significant loci explain roughly 3.4% of the variation in schizophrenia liability using the Lee et al. (2012) coefficient of determination. Using this estimate as a baseline, we select three choices for *θ*^2^: 0.02, 0.034 and 0.05. Our findings are consistent with a modest positive effect of schizophrenia liability on the odds of cannabis use. As shown in Figure 4, a one standard deviation increase in schizophrenia liability corresponds to a 1.15-1.26 increase in the odds of cannabis use, with 95% confidence interval range of 1.10-1.44. It is important not to directly compare these estimates with the original estimates: the two are not on the same scale. We must interpret the estimates of Table 4 in terms of standard deviation increases in schizophrenia liability.

**Figure 4:**
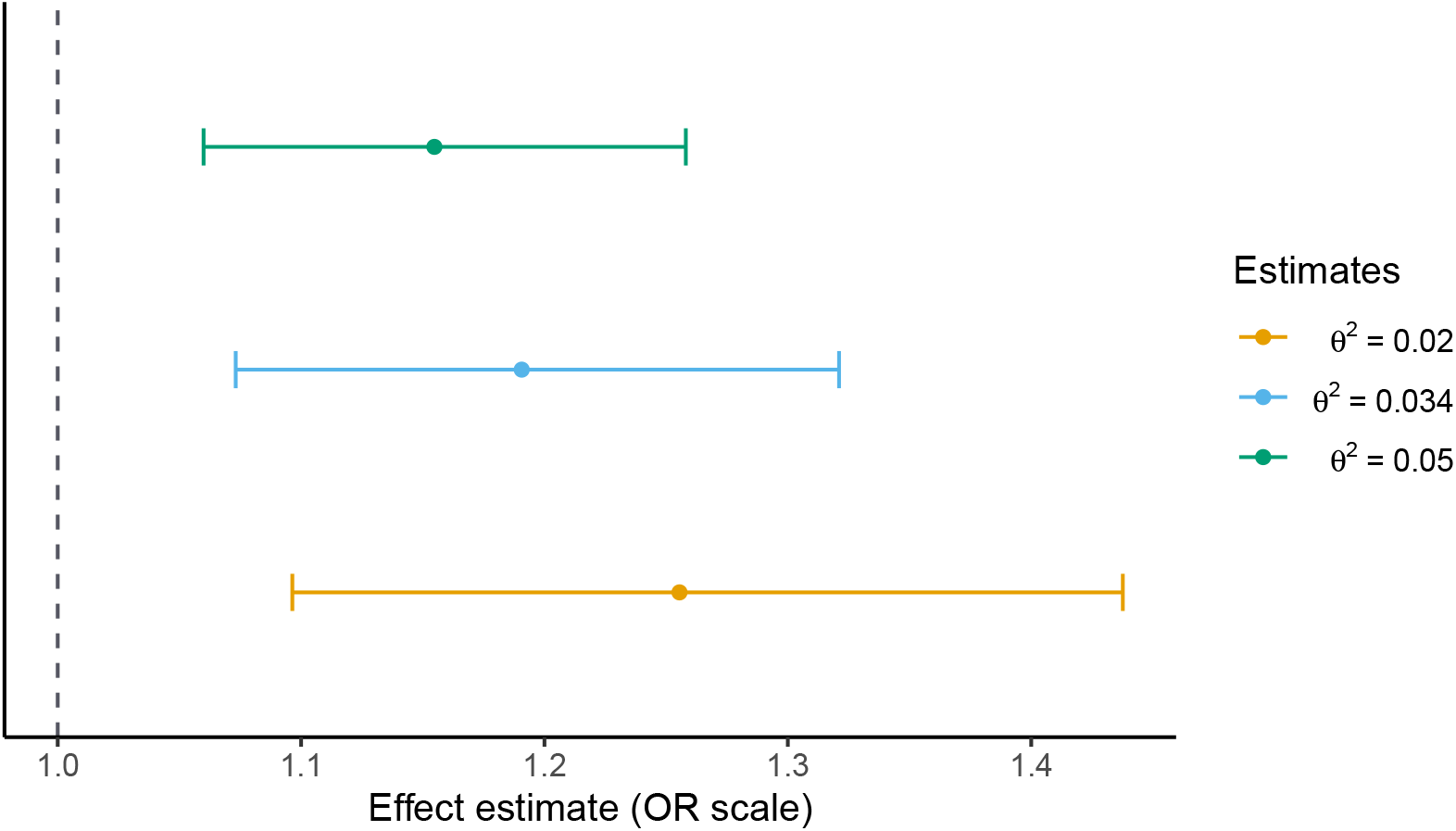
Effect of schizophrenia liability on risk of ever using cannabis for several choices of sensitivity parameter *θ*^2^. 95% confidence intervals are estimated as in Section C of the Appendix.

**Box 2:** Re-analysis of Richardson et al. (2020)

Richardson et al. (2020) performs two-sample Mendelian randomisation analysis of child and adult BMI on risk of several diseases: coronary artery disease, type 2 diabetes, breast cancer and prostate cancer. The instrument-exposure relationship is estimated in the UK Biobank cohort. However, child BMI is not measured directly in UK Biobank, instead, there is a measure of self-reported adiposity in three discrete categories (‘thinner’, ‘plumper’ or ‘about average’). In this context, the latent exposure is child BMI and the self-report measure is a coarsening of child BMI. Since the genetic instruments will act on child BMI directly, the exclusion restriction is likely to be violated.

Therefore, we apply the latent variable method of Section 3.2 to this data. We re-analyse the original univariable effect of child BMI on risk of type 2 diabetes (OR 2.32, 95% CI 1.76-3.05), coronary artery disease (1.49, 1.33-1.68) and breast cancer (0.59, 0.50-0.71).

We apply the two-sample generalisation of the inverse-variance weighted estimator of Section 4.4, estimating the instrument-exposure relationship in UK Biobank using an ordered probit model and the instrument-outcome relationships using the MR-Base platform (Hemani et al., 2018). We choose three values for *θ*^2^ based on a large GWAS of adult BMI: 0.01, 0.02 and 0.05 (Locke et al., 2015). The genetic share of child BMI is estimated using an ordered probit model and standard errors are calculated using the formula in Section C of the Appendix.

Figure 5 shows our results for three of the diseases analysed in the paper. Our estimates are in the same direction as the original estimates, which is expected, however, the interpretation of the magnitudes is different. For example, the original paper estimates that a per-category increase in self-reported child adiposity corresponds to an increase in the odds of coronary artery disease of 1.49 (95% CI 1.33-1.68), which could be inflated due to violation of the exclusion restriction. For *θ*^2^ = 0.02, we estimate that a one standard deviation increase in child BMI corresponds to an increase in the odds of coronary artery disease of 1.13 (95% CI 0.99-1.28). It is difficult to directly compare the two sets of estimates since the exposures are different, however, our estimate is suggestive of a modest effect of child BMI on the risk of coronary artery disease.

**Figure 5:**
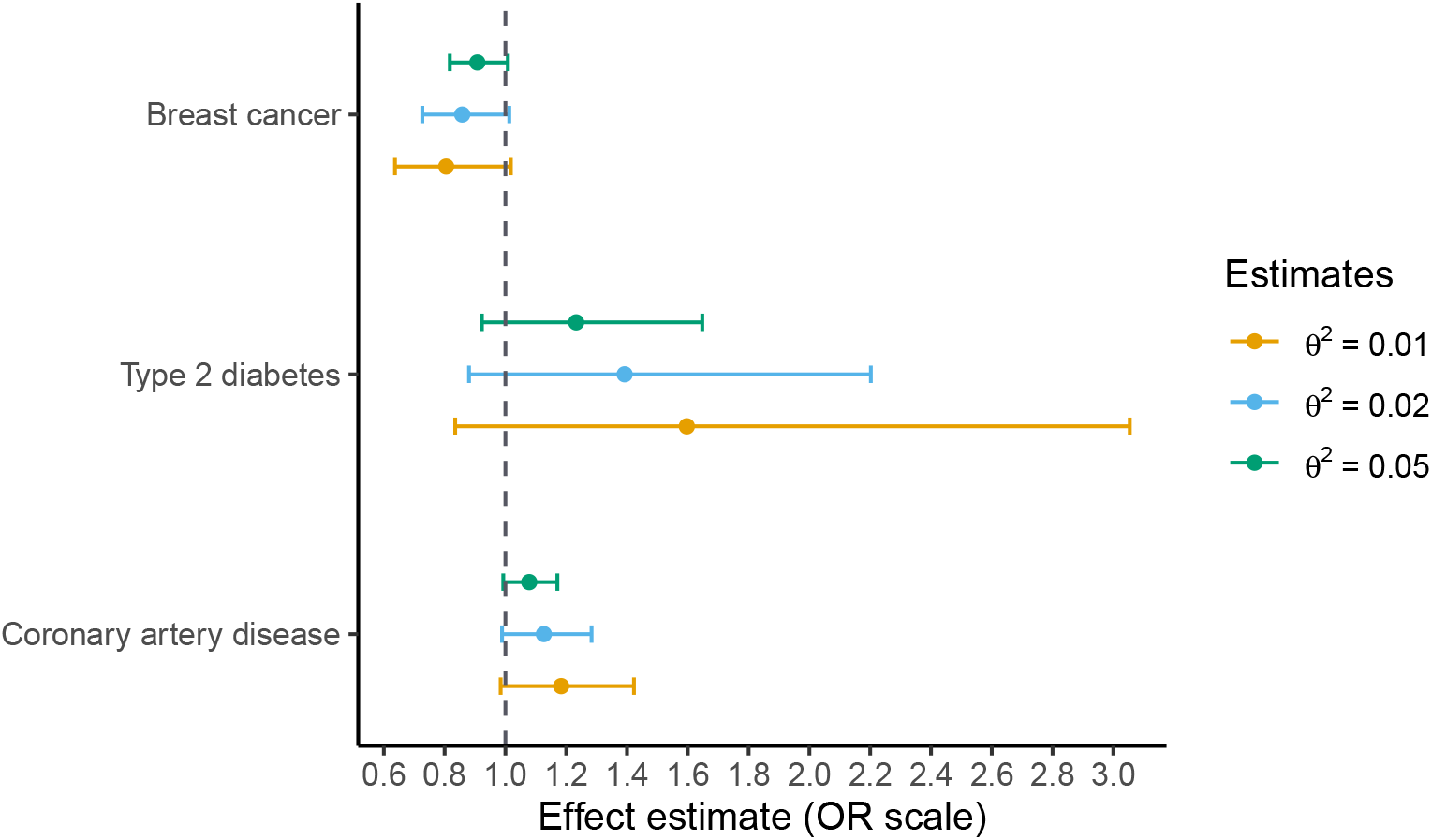
Effect of childhood BMI on risk of several diseases for several choices of sensitivity parameter *θ*^2^. 95% confidence intervals are estimated as in Section C of the Appendix.

## Data Availability

A replication kit for the analyses presented in this paper is available on GitHub. Access to the full genetic and phenotype data from UK Biobank waves 1 and 2 is required to replicate all of the analyses. Information regarding access to UK Biobank can be found on their website.

https://github.com/matt-tudball/mrlat_replication

https://www.ukbiobank.ac.uk/

## Acknowledgements

This research has been conducted using the UK Biobank Resource. UK Biobank received ethical approval from the Research Ethics Committee (REC reference for UK Biobank is 11/NW/0382). This research was approved as part of application 16391. This research was funded in whole, or in part, by the Wellcome Trust [220067/Z/20/Z]. For the purpose of Open Access, the author has applied a CC BY public copyright licence to any Author Accepted Manuscript version arising from this submission.

## Author contribution statement

GDS and MJT conceived the idea. MJT designed the method and performed the analyses. GDS, KT, QZ and RAH supervised the project. All authors contributed to the main ideas and the writing of the manuscript.

## Data availability statement

A replication kit for the analyses presented in this paper can be obtained from https://github.com/matt-tudball/mrlat_replication. Access to the full genetic and phenotype data from UK Biobank waves 1 and 2 is required to replicate Figures 3 and 5. UK Biobank is an open access resource available to bona fide scientists who are undertaking health-related research that is in the public good. Information regarding access to UK Biobank can be found at https://www.ukbiobank.ac.uk.

## Appendix

### A Importance of the identifying assumptions

Assumptions 1 and 3 require that the latent exposure and measurement are related by a linear single index model. This assumption imposes considerable structure on the relationship between the two. To see why this assumption is necessary for identification, consider the more general model *L* = *G* − *V* = *ν*(*Z, X*) − *V*, where *ν*(*·*) is some continuous function. *D* is invariant to any monotone transformation *t*(*·*) in the sense that

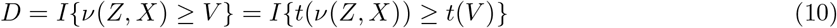

One such monotone transformation we can take is *t*(*·*) = *F*_*V*_ (*·*), where *F*_*V*_ (*·*) is the cumulative distribution of *V*, such that

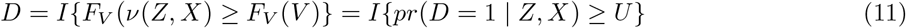

where *U ∼ Unif* (0, 1). This means that the observable data distribution *f* (*Z, X, D, Y*) is consistent with any monotone transformation of *ν*(*Z, X*), including *pr*(*D* = 1 | *Z, X*) itself. By imposing the structural assumption that *G* = *ν*(*Z, X*) = *µ* + *αZ* + *γ*^*1*^*X*, we reduce the class of models that the observed data distribution is consistent with to *ν*(*Z, X*) that proportional to *G*. This allows us to separate *G*, which is linear in parameters, from the non-linear link function. In the absence of this linear index assumption, this separation does not occur. This approach to identification is within the class of ‘identification by functional form’ methods described in Lewbel (2019), which provides an overview of this class of methods and discusses their limitations. Section B provides some simulation results when other assumptions fail, namely, correct specification of the link function and independence between the threshold and *Z* and *X*, both of which can introduce considerable bias. Bias from misspecification of the link function can be ameliorated by using more flexible semi-parametric binary outcome estimators (Ichimura, 1993; Klein & Spady, 1993). Independence of the threshold from *Z* and *X* is a reasonable assumption when these are genetic factors and *D* is disease diagnosis or when *D* is a deterministic categorisation of the latent exposure (i.e. splitting BMI into obesity status).

### B Simulating violations of the identifying assumptions

In this section, we present some simulation results which violate the identifying assumptions stated in Section 2. Our data generating process is as follows:

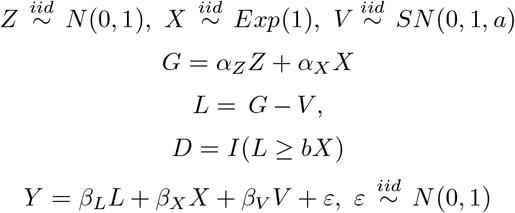

We set parameters (*α*_*Z*_, *α*_*X*_, *β*_*L*_,) equal to 1, (*β*_*X*_, *β*_*V*_) equal to 0.2, normalise *Z, X* and *V* to have mean 0 and normalise the variances as follows: *var* (*Z*) = 2*θ*^2^ /5, *var* (*X*) = 3*θ*^2^ /5 and *var*(*V*) = 1 − *θ*^2^, where *θ*^2^ = 0.1, meaning that *σ*_*L*_ = 1. *SN* (0, 1, *a*) denotes the skew normal distribution with skewness parameter *a*. We vary the skewness parameter over a range of values in Table 1. When *a* = 0, this is equivalent to the standard normal distribution, meaning that the probit link will be correctly specified. As *V* becomes more skewed, the bias increases. This bias can be ameliorated with semi-parametric methods for binary outcomes (Ichimura, 1993; Klein & Spady, 1993).

**Table 1:** Ratio of estimated to true *β*_*L*_ with link function misspecification

| Choice of link function | Skewness parameter $a$ | | | | | |
| --- | --- | --- | --- | --- | --- | --- |
|  | 0 | 1 | 2 | 3 | 4 | 5 |
| Logistic | 1.01<br>[1.01, 1.02] | 1.02<br>[1.01, 1.03] | 1.03<br>[1.03, 1.04] | 1.05<br>[1.04, 1.06] | 1.06<br>[1.05, 1.07] | 1.07<br>[1.06, 1.07] |
| Probit | 1.01<br>[1.00, 1.02] | 1.02<br>[1.01, 1.03] | 1.03<br>[1.02, 1.04] | 1.05<br>[1.04, 1.06] | 1.06<br>[1.05, 1.07] | 1.07<br>[1.06, 1.08] |
| Semi-parametric<sup>a</sup> | 1.00<br>[0.99, 1.00] | 1.00<br>[0.99, 1.01] | 1.00<br>[0.99, 1.01] | 1.00<br>[1.00, 1.01] | 1.01<br>[1.00, 1.01] | 1.01<br>[1.00, 1.02] |

<sup>a</sup>Klein and Spady estimator; mean over 1,000 draws; $N = 2,500$ ; $b = 0$ ; 95% Monte Carlo confidence intervals.

Another assumption that can be violated is independence between the threshold of *D* and the observed variables *Z* and *X*. This dependence is captured by the parameter *b*. Since *α*_*X*_ = 1, *b* can be interpreted as the relative contribution of *X* to the threshold compared to the latent exposure *L* (e.g. *b* = 0.5 means that *X* contributes half as much to the threshold as to the latent exposure). In Table 2, we vary the parameter *b* over a range of values and report the resulting bias. Despite the link function being correctly specified, there is significant bias from dependence in the threshold, which is roughly equal to *b* (e.g. when *b* = 0.5, relative bias in *β*_*L*_ is roughly 50%). Unlike misspecification of the link function, semi-parametric techniques cannot correct this bias. When *X* determines the threshold value, we cannot separately identify *G* in this framework. This simulation also suggests that threshold dependence may be bigger concern in this approach than misspecification of the link function.

**Table 2:** Ratio of estimated to true *β*_*L*_ with threshold dependence

| Choice of link function | Dependence parameter $b$ | | | | |
| --- | --- | --- | --- | --- | --- |
|  | 0 | 0.1 | 0.25 | 0.5 | 1 |
| Logistic | 1.01<br>[1.01, 1.02] | 1.08<br>[1.07, 1.08] | 1.17<br>[1.16, 1.18] | 1.34<br>[1.33, 1.36] | 1.71<br>[1.69, 1.72] |
| Probit | 1.01<br>[1, 1.02] | 1.07<br>[1.06, 1.08] | 1.17<br>[1.16, 1.18] | 1.33<br>[1.32, 1.34] | 1.69<br>[1.68, 1.70] |
| Semi-parametric<sup>a</sup> | 1.00<br>[0.99, 1] | 1.05<br>[1.05, 1.06] | 1.15<br>[1.14, 1.16] | 1.30<br>[1.29, 1.31] | 1.67<br>[1.66, 1.69] |

<sup>a</sup>Klein and Spady estimator; mean over 1,000 draws; $N = 2,500$ ; $a = 0$ ; 95% Monte Carlo confidence intervals.

### C Two-sample estimator and variance derivation

We begin by deriving equation (9). For some instrument *Z*_*ki*_ in 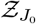, the estimand *β*_*G*_ can be written as

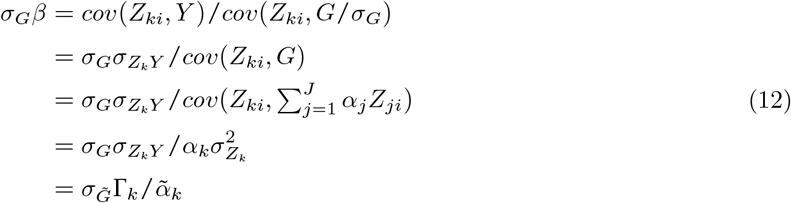

which we can estimate from GWAS summary data. We can use inverse-variance weighting to ‘meta-analyse’ over these estimates for each *Z*_*ki*_ in 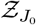, which recovers the estimator (9). Denote 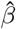 as the inverse-variance weighted estimator for *σ*_*V*_ *β*, then our two-sample estimator can be written as

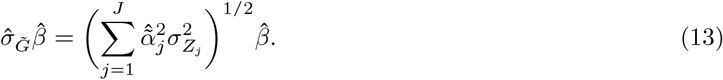

If we make the common assumption that 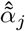 has negligible uncertainty 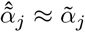, then we can write an estimator for the variance of (13) as

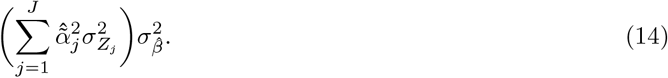

## Notes

This research was generously supported by the Wellcome Trust [220067/Z/20/Z]. The authors have no conflicts of interest to disclose.

### Competing Interest Statement

The authors have declared no competing interest.

### Author Declarations

UK Biobank has obtained Research Tissue Bank (RTB) approval from its ethics committee. The Research Ethics Committee reference for UK Biobank is 11/NW/0382.

### Summary of Updates

Some changes in terminology (introduces the term 'coarsened'), minor updates to figure and funding statement.

